# Creating an evidence-based economic model for prefilled parenteral medication delivery in the hospital setting

**DOI:** 10.1101/2022.11.04.22281902

**Authors:** Job F.H. Eijsink, Mia Weiss, Ashley Taneja, Tyler Edwards, Haymen Girgis, Betsy J. Lahue, Kristen A. Cribbs, Emily W. Acker, Maarten J. Postma

## Abstract

**Objectives:** Prefilled syringes (PFS) may offer clinical and economic advantages to conventional parenteral medication delivery methods (vials and ampoules). The benefits of converting from vials and ampoules to PFS have been elucidated in previous drug-specific economic models; however, these models have limited generalizability to different drugs, health care settings and other countries. This study aims to (1) present a comprehensive economic model to assess the impact of switching from vials to PFS delivery, and (2) illustrate the model’s utility by highlighting important features of shifting from vials to PFS through two case studies.

**Methods:** The economic model estimates the potential benefit of switching to PFS associated with four key outcomes: preventable adverse drug events (pADE), preparation time, unused drug, and cost of supplies. Model reference values were derived from existing peer-reviewed literature sources. The user inputs specific information related to the department, drug, and dose of interest can change reference values.

Two hypothetical case studies are presented to showcase model utility. The first concerns a cardiac intensive care unit in the United Kingdom administering 30 doses of 1mg/10mL atropine/day. The second concerns a COVID-19 intensive care unit in France that administers 30 doses of 10mg/25mL ephedrine/day.

**Results:** Total cost savings per hospital per year, associated with reductions in pADEs, unused drugs, drug cost and cost of supplies were £34,829 for the atropine example and €108,565 for the ephedrine example. Annual preparation time decreased by 371 and 234 hours in the atropine and ephedrine examples, respectively.

**Conclusions:** The model provides a generalizable framework with customizable inputs, giving hospitals a comprehensive view of the clinical and economic value of adopting PFS. Despite increased costs per dose with PFS, the hypothetical case studies showed notable reductions in medication preparation time and a net budget savings owing to fewer pADEs and reduced drug wastage.

**KEY MESSAGES:**

- Globally, most parenteral medications are supplied via injection with medication dispensed from vials and ampoules, despite evidence that such formats result in unused drug, increase risk of preventable adverse drug events, significant hospital staff time to prepare and use of extra supplies.
- Prefilled syringes address the shortcomings of these conventional parenteral medication delivery methods, with benefits for patients, healthcare delivery systems, and hospitals.
- A novel economic model was developed to estimate the holistic budget impact of switching from vials and/or ampoules to prefilled syringe medication delivery formats for acute care hospital settings.
- Results from two hypothetical case studies illustrate an overall cost offset despite higher prices of ready-to-administer formats with prefilled syringes compared to conventional delivery methods.

## INTRODUCTION

Throughout Europe, parenteral medication is predominantly supplied in vials and ampoules (referred to as conventional methods)^1^ despite documented limitations that negatively impact patients and healthcare systems.^2-7^ Ready-to-administer medication formats, including prefilled syringes (PFS), have the potential to redress conventional delivery shortcomings, yet only 2% of acute liquid injectable small molecule drugs ≤50mL are currently delivered in such formats, suggesting ample opportunity for improvement.^8^ Understanding the economic benefits of PFS versus conventional methods can support broader uptake of this modality.

Conventional methods are associated with many humanistic and economic implications. In fast-paced areas of the hospital where medications must be delivered quickly (e.g., Intensive Care Unit (ICU) or emergency departments), the risk of medication errors is higher.^2-4^ A German medical record-based study found that on average each adverse drug event (ADE) results in an additional cost of €970 for the health system and may be associated with downstream patient consequences.^5^ Additionally, dose preparation with conventional methods is complex and time-consuming, placing a substantial burden on healthcare professionals and hospital department resources.^6^ Furthermore, conventionally prepared doses are frequently discarded when narrow administration windows or expiration times are not met, resulting in unused drugs and supplies. In fact, one study estimated that 85% of all atropine doses prepared in operating rooms are discarded.^7^ As shown above, not only can conventional methods lead to avoidable adverse events for patients, but these delivery methods also yield significant inefficiencies and misallocation of resources, with extra costs for healthcare systems.

PFS provides a convenient solution to many of the shortcomings of conventional methods, with benefits for stakeholders, including healthcare delivery systems, hospitals, and patients. Since the medication is in a ready-to-administer format, PFS utilization reduces the number of steps required to deliver medication, which translates into a reduction in healthcare professional time spent preparing the injection.^6^ Additionally, eliminating preparation steps, such as drawing medication from vial and switching between the aspiration needle to the injection needle can decrease the contamination risk.^6,9,10^ Utilizing PFS has fewer steps required to deliver medications, thereby minimizing the risk of medication errors associated with conventional methods, including syringe preparation and the potential cascade of preventable ADEs (pADEs) that may follow. In fact, one study demonstrated that dosing errors were 17 times less likely in PFS versus conventional methods.^11^ Furthermore, while drugs prepared with conventional drug preparation methods have a limited period for sterile administration, PFS doses remain sterile under correct storage conditions until they are administered and therefore are less likely to be wasted.^10,12,13^

Prior economic models have taken a specific view of the budget impact for an individual drug, hospital, or outcome; however, new evidence is emerging to add potential cost savings of switching from vials to PFS across different hospitals and healthcare delivery systems. ^7,14^ To facilitate a more comprehensive view of the economic impact of switching to PFS, an economic model incorporating data on pADEs, unused drugs, preparation time, and cost of supplies was developed for use across various hospital departments, drugs, and countries. Through the presentation of two case studies, this study aims to (1) present the holistic nature of the economic model, and (2) illustrate the model’s utility by highlighting important and distinct features and impacts of switching from conventional methods to PFS.

## METHODS

### Model Development

The economic model was developed through a multi-step process with adherence to the International Society for Pharmacoeconomics and Outcomes Research (ISPOR) guidelines (Supplementary Information Fiigure 1) for budget impact analyses. Targeted literature reviews informed the model framework. The initial framework and parameters were reviewed by three advisory boards of pharmacists, doctors, and hospital administrators in France, Germany, and the United Kingdom (UK). The advisory boards discussed and selected the four most pertinent parameters for inclusion in the model. As research continues, the model will be further refined based on the most current literature and findings.

### Model Structure

The goal of the model is to provide insight into the annual impact of shifting from drug delivered in conventional methods (vials and ampoules) to PFS from a hospital perspective across four outcome areas (Figure 1). The Excel-based economic model includes five user-facing worksheets (Overview, Institution Overview, Input, Results, and References). Model costs and prices are converted to specific country prices via purchasing power parities (PPP). The costs of equipment for PFS are included in the PFS cost per dose. The model allows for inserting institution- and drug-specific inputs, such as cost per vial unit dose, cost per PFS unit dose, and number of doses administered per day.

**Figure 1.**
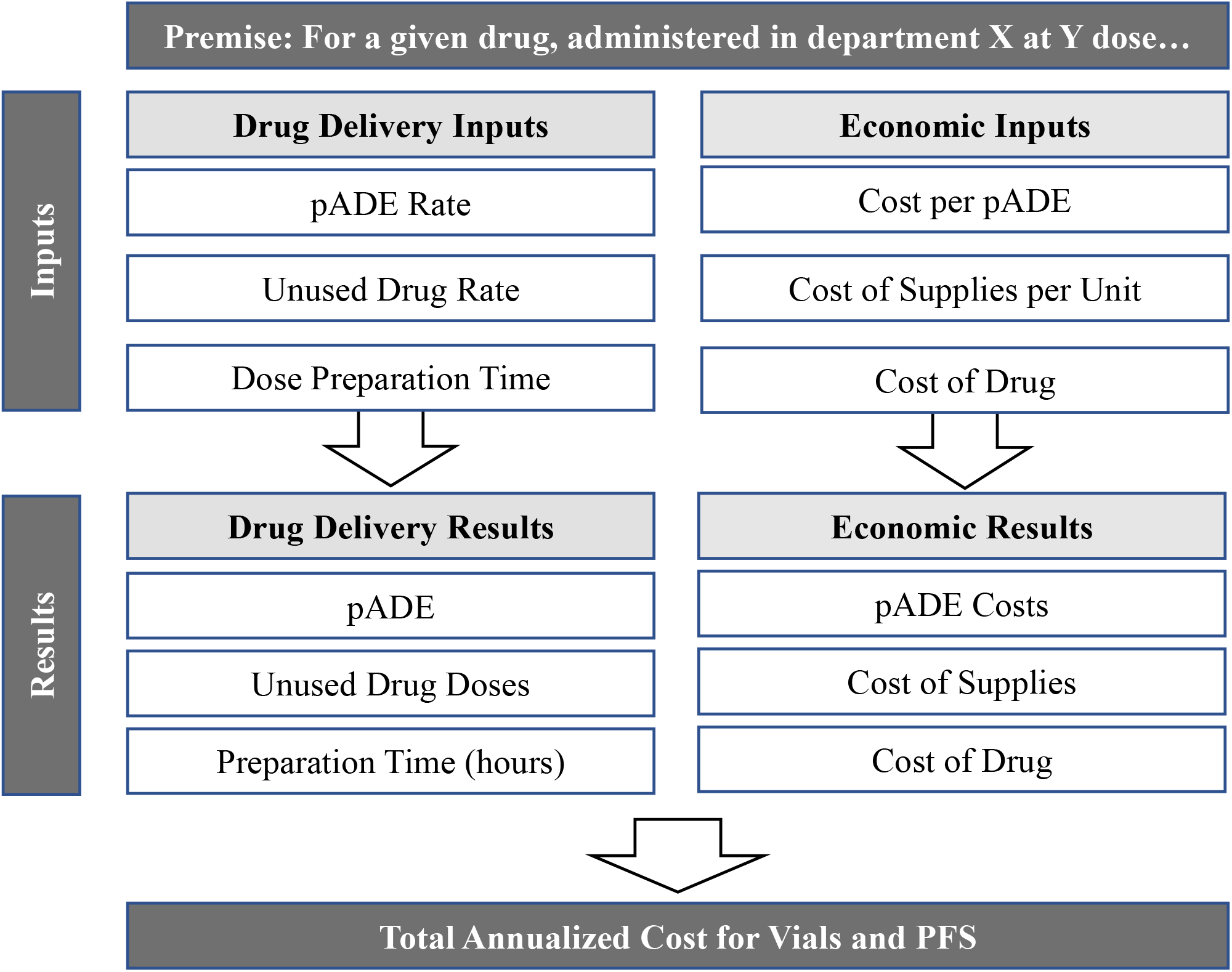
Economic Model Structure and Flow.

In addition to the cost of the drug, the model includes calculations for four main parameters: pADEs, unused drug, dose preparation time, and supplies per unit injection. The model input values are based on peer-review literature sources that were chosen to reflect the situation and drug of interest most accurately.

pADEs are a subset of adverse drug events that result from medication errors and can cause patient harm. The model reference pADE rates are 1.39 (51 pADEs recorded per 3,671 medication administrations) and 0.73 per 100 dose administrations for vials and PFS, respectively, and were derived from a United States (US) study of perioperative medication errors.^15^ The PFS pADE rate is calculated assuming that PFS introduction eliminates dosing and labeling errors (47.1% of all errors) and that all error types are equally likely to result in a pADE. The pADE rate can be changed in the model as requested for the specific analysis. A German medical record-based study showed that each ADE results in an incremental average direct hospital treatment cost of €970.^5^ The actual costs per ADE will vary from country to country due to conversion via PPP.

Unused drug is described as drug that is prepared but not used and therefore must be discarded.^7^ The model allows the user to either enter the percent of prepped doses unused per day or enter the total number of doses prepped per day and the model will calculate the number of unused doses per day. Additionally, the user may select to use reference values that vary by drug and drug dose for vials^12^ and a reference value of 3% for PFS.^10^ However, unused drug levels associated with conventional method preparation are highly dependent on the drug and type of hospital setting, therefore, reference values should only be used when institution level values are unknown.

Dose preparation time is defined as the total time it takes for hospital staff to prepare a single dose of medication.^6^ The preparation time assumptions are based on a time and motion study from two Danish hospitals and assumes 40.3 seconds per vial and 16.9 seconds per PFS.^6^ The model user may choose to select from additional options.^6,9,11,16^ Preparation time does not factor in monetarily to the cost calculations of the model.

Finally, the standard supplies included per unit injection costs of gloves, needles, syringes, and alcohol swabs.

### Case Studies

To showcase the robustness and utility of the economic model, two hypothetical case study analyses were conducted. Table 1 notes the assumptions and reference values for each case study. To ensure examples are reflective of current situations in hospitals in the UK and France, the assumptions and reference values utilized are based on local expert opinion and peer-reviewed literature. Drug costs are based on country-level IQVIA data for list prices from 2019-2021.^8^ The costs per dose were converted from United States Dollars ($) to Great British Pounds (£) or euros (€) for case studies 1 and 2, respectively.^8^

**Table 1.**
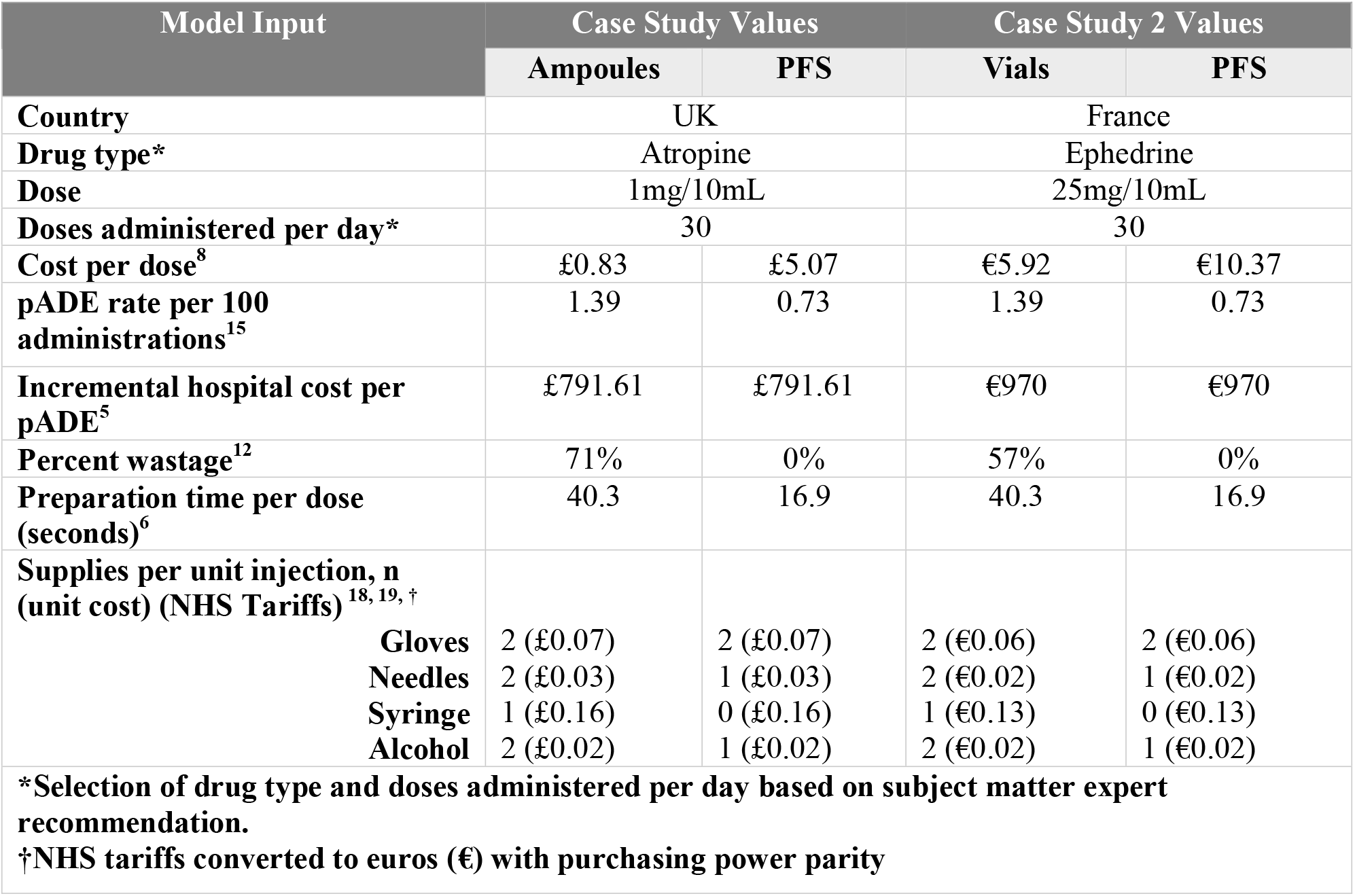
Economic Model Input: Case Study Values and References for Primary Analysis

### Case Study 1

Case study 1 takes place in a Cardiac Intensive Care Unit (CICU) in the UK that administers 30 doses of 1mg/10mL atropine per day. In the CICU setting, atropine is frequently used as a first-line therapy for symptomatic bradycardia, as well as a pre- and post-intubation medication. In this example, atropine doses from ampoules cost £0.83 per dose, and PFS format doses cost £5.07 per dose.^8^ The incremental cost of a pADE to a hospital system was £791.61.^5^ Unused drug levels for prepared doses were set at 71% and 0% for vials and PFS, respectively.^12^

### Case Study 2

Case study 2 takes place in an ICU that has been converted to a COVID-19 unit in France. The drug of interest is ephedrine – a vasopressor commonly used in operating rooms (ORs) and ICUs.^12^ This ICU uses 30 doses of 25mg/10mL ephedrine daily. The cost per vial dose is €5.92 and the cost per PFS dose is €10.37.^8^ The incremental cost of a pADE was €970.^5^ Unused drug levels were set at 57% and 0% for vials and PFS, respectively.^12^

### Sensitivity Analyses

Sensitivity analyses were conducted for both case studies using alternative references for drug waste to showcase model sensitivity. Case study 1 was repeated with the assumption that 85% and 0% of atropine doses were not used for vials and PFS, respectively.^7^ All other model parameters remained the same. Case study 2 was repeated with unused drug levels set at 74% and 0% for vials and PFS, respectively.^10^ All other model parameters remained the same (See Supplementary Information Taable 1).

## RESULTS

Key results from each case study are described below, with graphical depictions in Figures 2 and 3. Complete results are presented in Table 2.

**Table 2.**
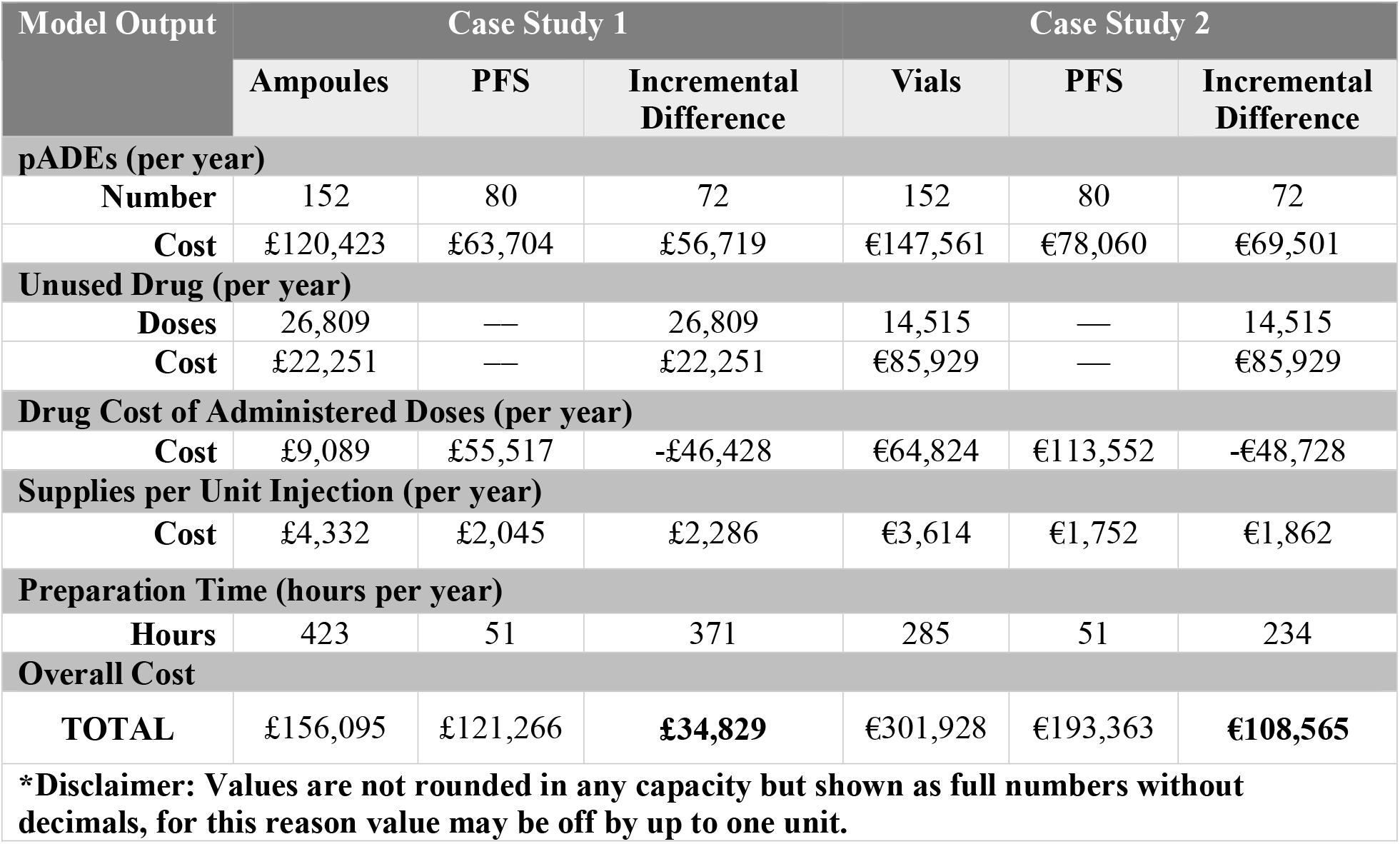
Economic Model Output: Primary Analysis Case Study Results

| Model Output | Case Study 1 |  |  | Case Study 2 |  |  |
| --- | --- | --- | --- | --- | --- | --- |
|  | Ampoules | PFS | Incremental Difference | Vials | PFS | Incremental Difference |
| **pADEs (per year)** |  |  |  |  |  |  |
| Number | 152 | 80 | 72 | 152 | 80 | 72 |
| Cost | £120,423 | £63,704 | £56,719 | €147,561 | €78,060 | €69,501 |
| **Unused Drug (per year)** |  |  |  |  |  |  |
| Doses | 26,809 | — | 26,809 | 14,515 | — | 14,515 |
| Cost | £22,251 | — | £22,251 | €85,929 | — | €85,929 |
| **Drug Cost of Administered Doses (per year)** |  |  |  |  |  |  |
| Cost | £9,089 | £55,517 | -£46,428 | €64,824 | €113,552 | -€48,728 |
| **Supplies per Unit Injection (per year)** |  |  |  |  |  |  |
| Cost | £4,332 | £2,045 | £2,286 | €3,614 | €1,752 | €1,862 |
| **Preparation Time (hours per year)** |  |  |  |  |  |  |
| Hours | 423 | 51 | 371 | 285 | 51 | 234 |
| **Overall Cost** |  |  |  |  |  |  |
| TOTAL | £156,095 | £121,266 | **£34,829** | €301,928 | €193,363 | **€108,565** |
| **\*Disclaimer: Values are not rounded in any capacity but shown as full numbers without decimals, for this reason value may be off by up to one unit.** |  |  |  |  |  |  |

**Figure 2.**
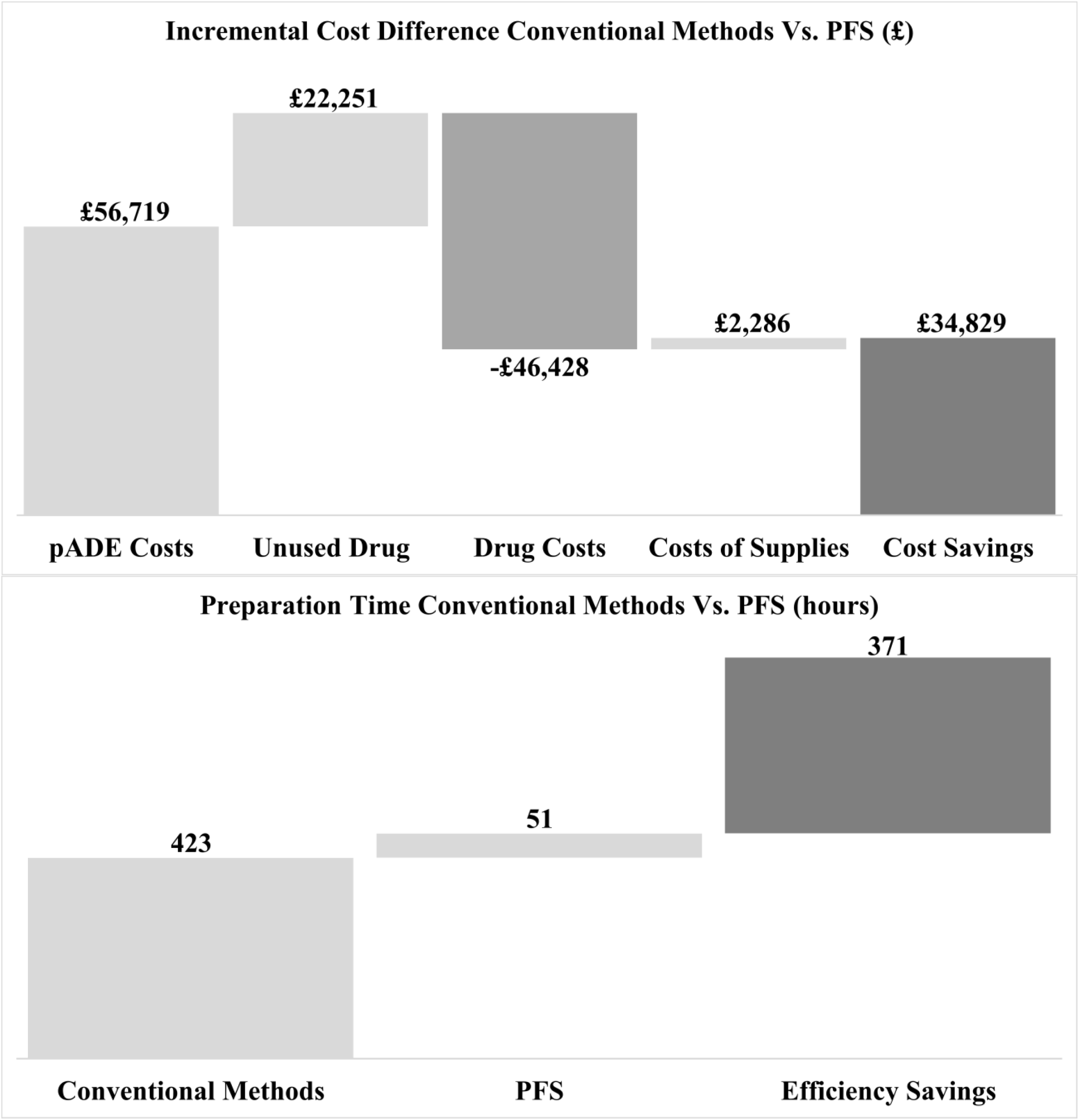
Graphical Depiction of Primary Case Study 1 Results**m**.

**Figure 3.**
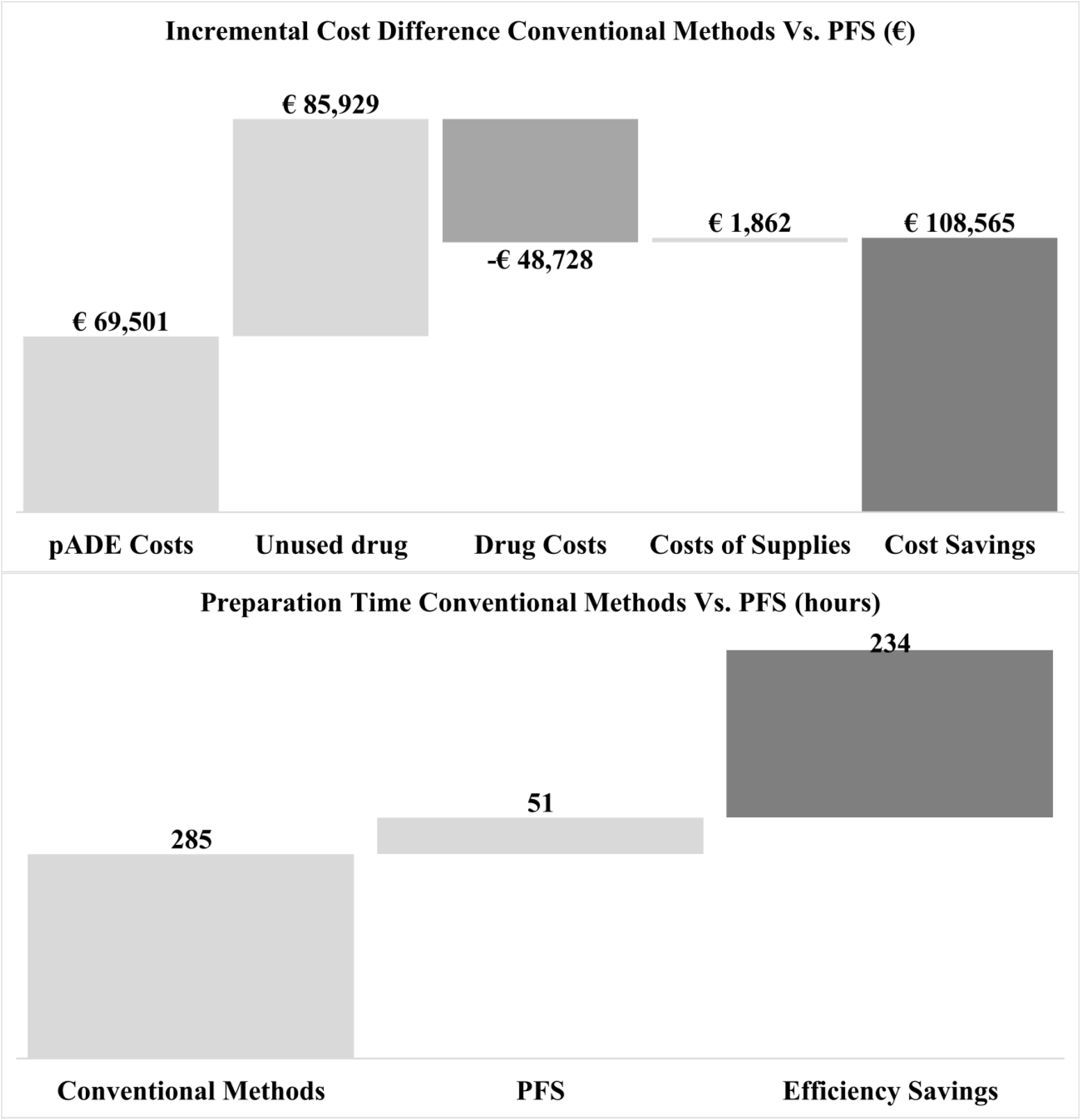
Graphical Depiction of Primary Case Study 2 Results.

### Case Study 1

Economic model results indicated that the overall cost impact for switching to PFS delivery in a hypothetical CICU in the UK administering 30 doses/day of atropine is £34,829 in savings per year. pADEs were reduced from 152 to 80 per year, and preparation time for hospital staff reduced from 423 to 51 hours per year.

### Case Study 2

The overall cost savings for a hypothetical COVID-19 ICU in France administering 30 doses/day of ephedrine is €108,565 per year based on the model results. pADEs were reduced from 152 to 80 per year, and preparation time for hospital staff reduced from 285 hours to 51 hours per year.

### Sensitivity Analyses

The sensitivity analysis for case study 1 (See Supplementary Information Taable 2 and Figure 2) revealed an overall cost savings of £64,079, with the cost of unused drug doses increasing to £51,502 (compared to £22,251 in the primary analysis). Preparation time savings was 766 hours per year compared to 251 hours per year in the primary analysis.

The sensitivity analysis for case study 2 (See Supplementary Information Taable 2 and Figure 3) showed an overall cost savings of €207,134, with the cost of unused drug doses increasing to €184,499 (compared to €85,929 in the primary analysis). Preparation time savings was 420 hours per year compared to 234 hours per year in the primary analysis.

## DISCUSSION

The economic model facilitates estimation of the budget impact of switching to PFS from conventional methods for institutions across four key outcomes associated with parenteral medication administration: pADE, preparation time, unused drug, and cost of supplies. Informed by peer-reviewed literature-based assumptions, hypothetical case studies and sensitivity analyses in two different settings with different drugs highlight the utility and versatility of the model, as well as the potential for hospital cost savings when switching from vials and ampoules to PFS. Despite increased costs per dose with PFS, the analysis in the case studies showed notable reductions in medication preparation time and a net budget savings owing to fewer pADEs and reduced drug waste.

Results from the case studies underscore that drug price significantly impacts model outcomes. The largest drivers of cost savings were found to be related to reductions in unused drug doses and pADEs in PFS versus conventional methods. However, exact prices for drugs cannot always be determined, which is why case studies use official prices without accounting for possible discounts. Despite lower unused drug levels seen in ephedrine compared to atropine, the costs of unused drug doses were higher in case study 2 compared to case study 1 due to higher costs per dose of ephedrine in vials. The cost difference between vial and PFS format will substantially impact the level of cost savings that could be achieved.

The sensitivity analyses conducted show how much results can change with a single manipulation of the model, showcasing the value of the tool in practical settings. Sensitivity analyses were conducted using varying levels of unused drug for the primary case studies. Other studies have revealed that different drugs are associated with different levels of unused drug.^7,10,12^ Results indicated that the economic model is sensitive to changes in unused drug levels, with implications for the total cost savings. The primary and sensitivity analyses for both case studies assumed zero waste for PFS; however, it should be acknowledged that there is a potential for discarded doses (e.g., if sterility is broken or if the PFS dose is left unrefrigerated for too long).

Notably, as demonstrated through the case studies, PFS drug administration is projected to nearly halve the estimated pADEs, which has critical implications for individual patient safety and related costs, including reducing excess hospitalizations and length of hospital stays.^5,17^ Ultimately, the case studies show that the higher upfront costs of PFS may be offset by reductions in pADE and unused drug, potentially leading to overall reduced costs.

Results revealed that preparation time savings were higher for atropine compared to ephedrine. One reason for higher preparation time savings is due to higher levels of unused drug doses in atropine. All prepared doses contribute to staff time, and the higher the levels of unused drugs, the more staff time is used preparing drugs that are ultimately wasted. Therefore, preparation time, for drug that is both used and unused, is higher for atropine compared to ephedrine. Similarly high preparation time savings would be expected for other high-acuity drugs, such as epinephrine and midazolam, which are often prepared in advance of administration and must frequently be discarded.^12^ For drugs similar to ephedrine that are commonly used and prepared in advance, the preparation time savings from shifting from vials to PFS are expected to be lower.

## Limitations

There are several limitations of the model and analysis. The complexity of switching from conventional methods to PFS is simplified in the economic model to include four main domains that were chosen based on availability of evidence and amenability to modeling. The underlying data that drives the model is specific but may not adequately reflect the individual setting of interest because it assumes that conditions are similar at the user site and the reference site. There is a particular scarcity of pADE rate data, so the model uses a US-based source. Given variation in dose preparation time in the literature, the model utilizes a conservative lower-end estimate, which may lead to an underestimated result. The effect of underestimation is minimized due to the model examining changes before and after switching from conventional methods to PFS methods; however, the difference between conventional methods preparation and PFS preparation may be significantly greater than the model estimates. Further, the outcomes modeled may not be applicable to all drug uses and may not reflect all benefits and costs of PFS. Finally, the model does not account for certain factors that may influence costs of switching from conventional methods to PFS from a global perspective, including microbial contamination risks, costs of sharps disposal, and storage costs and requirements for PFS.

## CONCLUSION

Throughout the COVID-19 pandemic, shortages of hospital staff, especially ICU nurses, have impacted the efficiency of care and overall health system burden. Challenges in healthcare delivery during the past two years highlight the importance of dissemination of existing innovations into new territory, including the adoption of PFS, to improve efficiency and patient safety for now and as we look to future challenges and additional potential pandemics.

Results from the two case studies reinforce that relevant cost savings can be realized across various drugs with differing use-cases, settings, and practice patterns when switching from vials and ampoules to PFS. The developed model shows important financial, clinical, and humanistic implications for various stakeholder groups, highlighting its utility for decision makers. While the examples included in this study were intended to mimic real-world acute care settings, future model users are encouraged to use individualized hospital or department data, where possible, to increase the accuracy of the model and relevance of findings.

## Supporting information

See Supplementary Information Table 2 and Figure 2

See Supplementary Information Table 2 and Figure 3

## Data Availability

All data produced in the present work are contained in the manuscript

## ACKNOWLEDGEMENTS AND AFFILIATIONS

- Alkemi LLC received consultancy fees from BD.
- The authors would like to thank Cecile Frolet for her guidance during the development of the publication.

## REFERENCES

1. Schaut RA, Weeks WP. Historical Review of Glasses Used for Parenteral Packaging. PDA J Pharm Sci Technol 2017;71(4):279–96. doi: 10.5731/pdajpst.2016.007377 [published Online First: 2017/02/16]

2. Keers RN, Williams SD, Cooke J, et al. Prevalence and nature of medication administration errors in health care settings: a systematic review of direct observational evidence. Ann Pharmacother 2013;47(2):237–56. doi: 10.1345/aph.1R147 [published Online First: 2013/02/07]

3. Berdot S, Gillaizeau F, Caruba T, et al. Drug administration errors in hospital inpatients: a systematic review. PLoS One 2013;8(6):e68856. doi: 10.1371/journal.pone.0068856 [published Online First: 2013/07/03]

4. Leendertse AJ, Egberts AC, Stoker LJ, et al. Frequency of and risk factors for preventable medication-related hospital admissions in the Netherlands. Arch Intern Med 2008;168(17):1890–6. doi: 10.1001/archinternmed.2008.3 [published Online First: 2008/09/24]

5. Rottenkolber D, Hasford J, Stausberg J. Costs of adverse drug events in German hospitals--a microcosting study. Value Health 2012;15(6):868–75. doi: 10.1016/j.jval.2012.05.007 [published Online First: 2012/09/25]

6. Subhi Y, Kjer B, Munch IC. Prefilled syringes for intravitreal injection reduce preparation time. Dan Med J 2016;63(4) [published Online First: 2016/04/02]

7. Benhamou D, Piriou V, De Vaumas C, et al. Ready-to-use pre-filled syringes of atropine for anaesthesia care in French hospitals - a budget impact analysis. Anaesth Crit Care Pain Med 2017;36(2):115–21. doi: 10.1016/j.accpm.2016.03.009 [published Online First: 2016/08/03]

8. IQVIA. Injectable drug universe data in IQVIA Smart MIDAS 2019, 2019-2021.

9. Souied E, Nghiem-Buffet S, Leteneux C, et al. Ranibizumab prefilled syringes: benefits of reduced syringe preparation times and less complex preparation procedures. Eur J Ophthalmol 2015;25(6):529–34. doi: 10.5301/ejo.5000629 [published Online First: 2015/06/06]

10. Atcheson CL, Spivack J, Williams R, et al. Preventable drug waste among anesthesia providers: opportunities for efficiency. J Clin Anesth 2016;30:24–32. doi: 10.1016/j.jclinane.2015.12.005 [published Online First: 2016/04/05]

11. Adapa RM, Mani V, Murray LJ, et al. Errors during the preparation of drug infusions: a randomized controlled trial. Br J Anaesth 2012;109(5):729–34. doi: 10.1093/bja/aes257 [published Online First: 2012/08/02]

12. Barbariol F, Deana C, Lucchese F, et al. Evaluation of Drug Wastage in the Operating Rooms and Intensive Care Units of a Regional Health Service. Anesth Analg 2021;132(5):1450–56. doi: 10.1213/ANE.0000000000005457 [published Online First: 2021/03/06]

13. Weinger MB. Drug wastage contributes significantly to the cost of routine anesthesia care. J Clin Anesth 2001;13(7):491–7. doi: 10.1016/s0952-8180(01)00317-8 [published Online First: 2001/11/13]

14. Larmene-Beld KHM, Spronk JT, Luttjeboer J, et al. A Cost Minimization Analysis of Ready-to-Administer Prefilled Sterilized Syringes in a Dutch Hospital. Clin Ther 2019;41(6):1139–50. doi: 10.1016/j.clinthera.2019.04.024 [published Online First: 2019/05/14]

15. Nanji KC, Patel A, Shaikh S, et al. Evaluation of Perioperative Medication Errors and Adverse Drug Events. Anesthesiology 2016;124(1):25–34. doi: 10.1097/ALN.0000000000000904 [published Online First: 2015/10/27]

16. Moreira ME, Hernandez C, Stevens AD, et al. Color-Coded Prefilled Medication Syringes Decrease Time to Delivery and Dosing Error in Simulated Emergency Department Pediatric Resuscitations. Ann Emerg Med 2015;66(2):97–106 e3. doi: 10.1016/j.annemergmed.2014.12.035 [published Online First: 2015/02/24]

17. Amelung S, Meid AD, Nafe M, et al. Association of preventable adverse drug events with inpatients’ length of stay-A propensity-matched cohort study. Int J Clin Pract 2017;71(10) doi: 10.1111/ijcp.12990 [published Online First: 2017/09/06]

18. National Health Service England And Wales: Electronic Drug Tariff. (2021, May). Retrieved May, 2021, from https://www. https://www.drugtariff.nhsbsa.nhs.uk/#/00803325-DC/DC00803258/Part%20IXA-Appliances.

19. Health care prices - brief - OECD. https://www.oecd.org/health/health-systems/Health-Care-Prices-Brief-May-2020.pdf. Accessed February 23, 2022.

