## Supplementary Information Table 1 for "Creating an evidence-based economic model for prefilled parenteral medication delivery in the hospital setting"

**Supplementary Material**

Supplemental Information Table 1: Economic Model Input Case Study Values and References for Sensitivity Analysis

| Model Input | Case Study Values | | Case Study 2 Values | |
|---|---|---|---|---|
| | Ampoules | PFS | Vials | PFS |
| **Country** | UK | | France | |
| **Drug type*** | Atropine | | Ephedrine | |
| **Dose** | 1mg/10mL | | 25mg/10mL | |
| **Doses administered per day*** | 30 | | 30 | |
| **Cost per dose[8]** | £0.83 | £5.07 | €5.92 | €10.37 |
| **pADE rate per 100 administrations[15]** | 1.39 | 0.73 | 1.39 | 0.73 |
| **Incremental hospital cost per pADE[5]** | £791.61 | £791.61 | €970 | €970 |
| **Percent wastage[7,10]** | 85% | 0% | 74% | 0% |
| **Preparation time per dose (seconds)[6]** | 40.3 | 16.9 | 40.3 | 16.9 |
| **Supplies per unit injection, n (unit cost) (NHS Tariffs) [18, 19, †]** | | | | |
| **Gloves** | 2 (£0.07) | 2 (£0.07) | 2 (€0.06) | 2 (€0.06) |
| **Needles** | 2 (£0.03) | 1 (£0.03) | 2 (€0.02) | 1 (€0.02) |
| **Syringe** | 1 (£0.16) | 0 (£0.16) | 1 (€0.13) | 0 (€0.13) |
| **Alcohol** | 2 (£0.02) | 1 (£0.02) | 2 (€0.02) | 1 (€0.02) |

*Selection of drug type and doses administered per day based on subject matter expert recommendation.
†NHS tariffs converted to euros (€) with purchasing power parity
