## Supplementary Information Table 2 for "Creating an evidence-based economic model for prefilled parenteral medication delivery in the hospital setting"

**Supplementary Material**

Supplemental Information Table 2. Economic Model Output: Sensitivity Analysis Case Study Results

| Model Output | Case Study 1 | | | Case Study 2 | | |
|---|---|---|---|---|---|---|
| | **Ampoules** | **PFS** | **Incremental Difference** | **Vials** | **PFS** | **Incremental Difference** |
| **pADEs (per year)** | | | | | | |
| Number | 152 | 80 | 72 | 152 | 80 | 72 |
| Cost | £120,423 | £63,704 | £56,719 | €147,561 | €78,060 | €69,501 |
| **Unused Drug (per year)** | | | | | | |
| Doses | 62,050 | — | 62,050 | 31,165 | — | 31,165 |
| Cost | £51,502 | — | £51,502 | €184,499 | — | €184,499 |
| **Drug Cost of Administered Doses (per year)** | | | | | | |
| Cost | £9,089 | £55,517 | -£46,428 | €64,824 | €113,552 | -€48,728 |
| **Supplies per Unit Injection (per year)** | | | | | | |
| Cost | £4,332 | £2,045 | £2,286 | €3,614 | €1,752 | €1,862 |
| **Preparation Time (hours per year)** | | | | | | |
| Hours | 817 | 51 | 766 | 471 | 51 | 420 |
| **Overall Cost** | | | | | | |
| **TOTAL** | £185,345 | £121,266 | **£64,079** | €400,498 | €193,363 | **€207,134** |
| *Disclaimer: Values are not rounded in any capacity but shown as full numbers without decimals, for this reason value may be off by up to one unit. | | | | | | |
