## Supplemental Information Figure 1 for "Creating an evidence-based economic model for prefilled parenteral medication delivery in the hospital setting"

### Supplementary Material

Supplemental Information Figure 1. Acknowledgement of ISPOR Guidelines for Budget Impact Analysis

**Table 1 – Aspects to be considered in the design of a budget impact analysis.**

- Features of the health care system
- Perspective
- Use and cost of current and new interventions
  - Eligible population
  - Current interventions
  - Uptake of new intervention and market effects
  - Off-label uses of the new intervention
  - Cost of the current or new intervention mix
- Impact on other costs
  - Condition-related costs
  - Indirect costs
- Time horizon
- Time dependencies and discounting
- Choice of computing framework
- Uncertainty and scenario analysis
- Validation
