## Supplementary material for "Creating an evidence-based economic model for prefilled parenteral medication delivery in the hospital setting": See Supplementary Information Table 2 and Figure 3

Supplemental Information Figure 3. Graphical Depiction of Sensitivity Case Study 2 Results

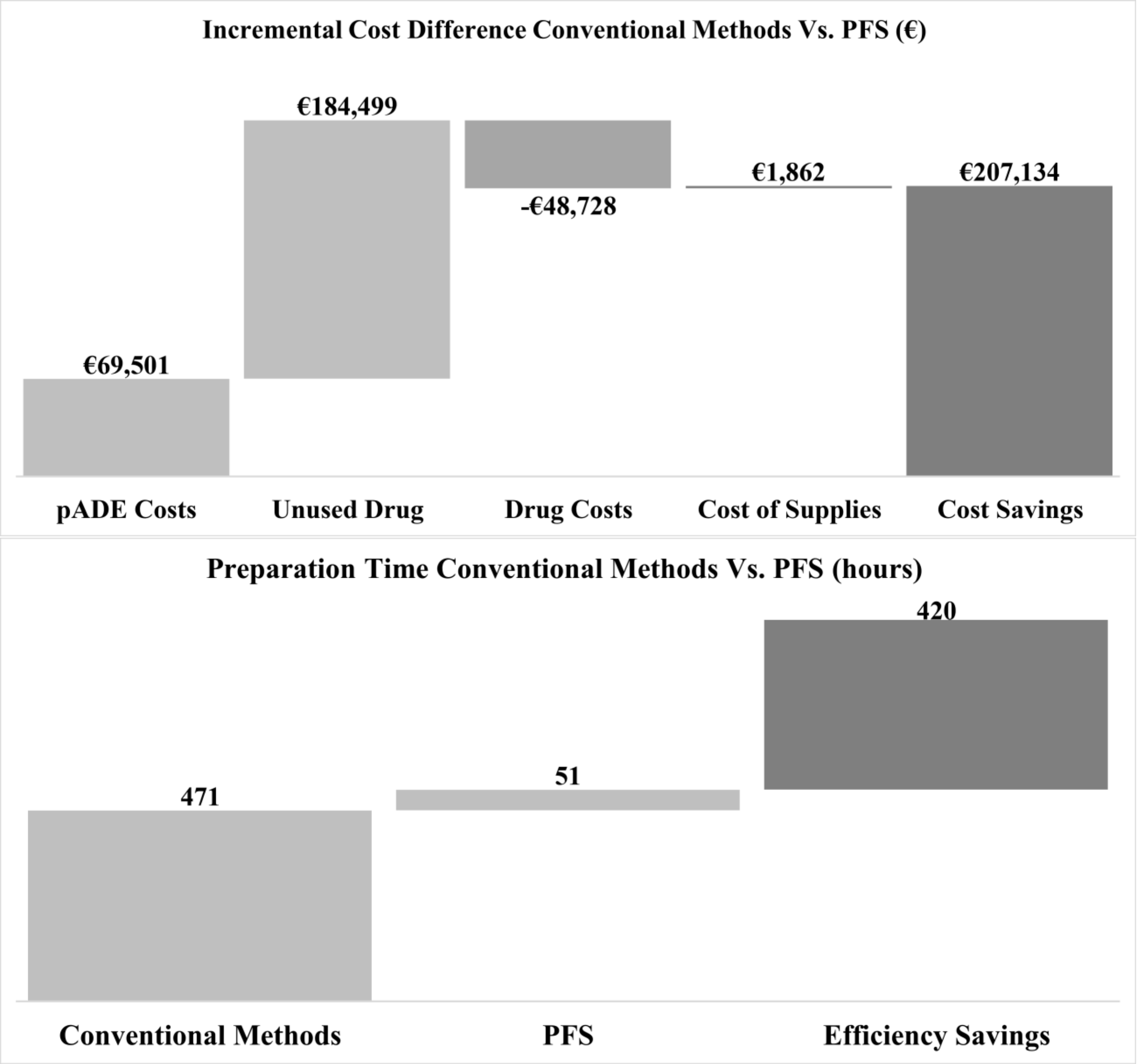
